# The dark side of the mood: structural and functional fronto-insular and cerebellar alterations classify major depression

**DOI:** 10.1101/2025.04.09.25325506

**Authors:** Rodolfo Rizzi, Khanitin Jornkokgoud, Parisa Ahmadi Ghomroudi, Massimo Stella, Alessandro Grecucci

## Abstract

Despite major depressive disorder (MDD) being the leading cause of disability worldwide, the exact characterization of its neural bases and the development of reliable biomarkers are still at an early stage. A possible solution lies in multimodal analysis approaches, which integrate cross-modal data to investigate the relationship between structural and functional network disruptions, potentially improving the accuracy of machine learning (ML) models for individual-level predictions. In this study, we employed a data fusion unsupervised ML method called transposed Independent Vector Analysis (tIVA) to investigate joint functional and structural brain networks to classify MDD. To this aim, the amplitude of low-frequency fluctuations (ALFF) and gray matter density (GMD) of 461 participants (MDD = 226, HC = 235) were taken into consideration. The analysis revealed a multimodal link between reduced functional activity in the cerebellum and structural deficits in subcortical regions (primarily including the anterior cingulate cortex (ACC) and insula) implicated in emotional regulation, highlighting how these structural and functional changes can mutually influence and reinforce each other. Moreover, enhanced functional activity was found in dorsomedial prefrontal areas of the default mode network (DMN), with concurrent reduced activity in dorsolateral prefrontal regions of the executive control network (CEN). Importantly, a Random Forest (RF) classifier, which identified the same areas as important classification features, achieved a 69.89% accuracy in distinguishing MDD patients from HC. These findings underscore the value of combining multimodal data-driven approaches to investigate the neural basis of MDD, possibly enhancing diagnostic precision and advancing precision psychiatry.

## Introduction

Major depressive disorder (MDD) is one of the most severe and prevalent neuropsychiatric disorders, affecting over 300 million people globally (World Health Organization, 2017). Prospective studies indicate that more than 30% of individuals may experience this mood disorder during their lifetime (Moffitt et al., 2010). MDD is characterized by a complex symptomatology manifestation that includes a persistent low or depressed mood, anhedonia, chronic fatigue, pervasive feelings of guilt or worthlessness, and recurring thoughts of death (Marx et al., 2023). Beyond MDD’s profound impact on patients’ personal quality of life, its societal burden is equally alarming. In 2019, the economic cost of this disorder was estimated at $333.7 billion in the United States alone (Greenberg et al., 2023), driven by healthcare costs, productivity losses, household-related expenses, presenteeism and absenteeism. Moreover, recent evidence suggests that this figure could worsen due to a significant increase in the number of MDD cases following the COVID-19 pandemic (Santomauro et al., 2021).

Although extensive research has been conducted on this mental disorder, its diagnosis and treatment remain challenging due to the absence of specific biomarkers (Fritz et al., 2017; Mousavian et al., 2021) and a lack of consensus in distinguishing clinical from subclinical manifestations (Marx et al., 2023). Indeed, it is well-documented how time-consuming and operational diagnostic criteria based on the Diagnostic and Statistical Manual of Mental Disorders Fifth Edition Text Revision and the International Classification of Diseases 11th Revision can be ineffective, resulting in misdiagnoses and significant delays in appropriate treatment (Fritz et al., 2017).

However, remarkable opportunities are emerging to develop personalized assessments and neurobiologically grounded models of pathological mental conditions, driven by the convergence of a growing number of open-access neuroscientific datasets and new artificial intelligence (AI) methodologies (Masdeu, 2011; Sui et al., 2012; Winter et al., 2022). In this direction, neuroimaging-based AI may enable a more objective characterization and explanation of psychiatric conditions and ultimately enhance clinical decision-making in early diagnosis, prognosis, and treatment (Eyre et al., 2016; Marx et al., 2023).

### Literature review and current limitations

Previous investigations on MDD have reported several structural and functional abnormalities, leading researchers to define this mental condition as a network-based disorder. Crucial for this conceptualization is the role of specific neural hubs, whose disruption can lead to maladaptive integration and segregation processes across the brain, contributing to this disorder’s cognitive and emotionally heterogeneous symptomatology (Menon, 2011; Chai et al., 2023). For instance, self-negative rumination has been associated with altered activity in prefrontal regions of the default mode network (DMN) (Hamilton et al., 2015), cognitive control deficits have been linked to reduced central executive network (CEN) activity (Menon, 2011), and attention and affective processing deficits have been related with structural and functional abnormalities in the salience network (SN) (Menon, 2011). Particularly, resting-state functional magnetic resonance imaging (rs-fMRI) studies consistently report functional abnormalities in regions such as the prefrontal cortex (PFC) (Jing et al., 2013), cerebellum (Wang et al., 2012; Gong et al., 2020), and limbic areas (Jing et al., 2013; Zhong et al., 2019). Among rs-fMRI indices, the amplitude of low-frequency fluctuations (ALFF) has been widely used to evaluate spontaneous brain activity related to blood oxygen level-dependent (BOLD) signals (Yan et al., 2017). This metric has proven to be reliable and stable in characterizing different neuropsychiatric conditions, such as schizophrenia (Athanassiou et al., 2022), post-traumatic stress disorder (Disner et al., 2018), MDD, and bipolar disorder (Gong et al., 2020). Specifically, ALFF patterns in MDD result to be increased in the superior frontal gyrus (SFG) and insula and decreased in the cerebellum and occipital cortex, reflecting disruptions in self-referential processing, affective regulation and sensory integration (Gong et al., 2020). Moreover, meta-analytic evidence highlights ALFF abnormalities and co-localized structural deficits in the subgenual anterior cingulate cortex (ACC), hippocampus, and amygdala, regions critical to emotion regulation and memory processing (Gray et al., 2020).

Additional evidence supporting the critical involvement of the PFC in the pathophysiology of depression comes from proton magnetic resonance spectroscopy (1H-MRS) studies, which revealed that altered glutamate/glutamine (Glx) levels may be implicated in this condition (Moriguchi et al., 2019). On the other hand, structural analyses consistently indicate reduced gray matter (GM) volume in the insula and cingulate cortex of MDD patients (Wise et al., 2017; Sha et al., 2019; Schmaal et al., 2020), while cross-disorder studies confirm transdiagnostic GM reductions in cortical and subcortical regions, particularly the orbitofrontal cortex (OFC) and anterior ACC (Yoshimura et al., 2014; Opel et al., 2020). Interestingly, a recent genome-wide meta-analysis has highlighted that these structural and functional abnormalities may be associated with 102 independent genetic variants linked to depression. These alterations contribute to disrupted synaptic structure and neurotransmission, particularly in the PFC and ACC, regions frequently implicated in MDD pathophysiology (Howard et al., 2019).

However, three significant limitations in the existing literature impede the identification of clear biomarkers for major depressive disorder (MDD) and contribute to the replication failures observed in depression research. The first limitation arises from the predominant use of mass univariate analyses in most prior studies, which are unable to account for the complex, network-based nature of brain-behavior relationships (Calhoun and Sui, 2016; Winter et al., 2022). This limitation may help explain the inconsistencies observed in earlier findings. The second limitation lies in the lack of generalizability in prior research, as predictive models were neither derived nor statistically validated (Wolfers et al., 2020; Guo et al., 2024). Consequently, previous findings are closely tied to the specific samples studied, without evaluating their applicability to other populations. The third limitation concerns the partial characterization of the neural bases of depression, which has predominantly relied on one modality-specific comparison per time. This approach fails to address the intricate and multifaceted nature of depression that affects several aspects of brain function and structure (Sui et al., 2023). Indeed, modern neuroscientific perspectives emphasize that neurological and psychiatric disorders can be better understood by examining both widespread voxel alterations and multiscale interactions across structural, functional and genetic levels (Biswal et al., 2010; van den Heuvel et al., 2019). Consequently, combining multimodal imaging techniques has shown the possibility to uncover robust and convergent differences in brain morphology and function by integrating cross-modal data (Meda et al., 2014; Gray et al., 2020; Sui et al., 2023). The final aim is to reveal how brain structure and function influence each other, thus identifying the physiological aspects that drive maladaptive cognition and behavior (Sui et al., 2012, 2014).

Previous investigations have attempted to overcome some key limitations. For instance, several studies have employed single modality ML classification methods primarily using rs-fMRI and sMRI features, with promising results (Gao et al., 2018; Bondi et al., 2023). However, most of these models were often trained and tested on small sample sizes and suffer from inconsistent methodologies, ultimately not providing significant advancement for translating neuroimaging biomarkers into clinical practice (Gao et al., 2018; Bondi et al., 2023). Other studies tried to address the limitations of single-modality studies using multimodal data fusion techniques. On the one hand, Qi et al. (Qi et al., 2018) used a multimodal canonical correlation analysis with a joint independent component analysis fusion-with-reference model (mCCAR+jICA) to combine fractional ALFF, gray matter volume, and fractional anisotropy and miR-132 blood levels to investigate the interrelationship between epigenetic factors and different brain features in unmedicated patients with MDD. The study found that miR-132 dysregulation was linked to reduced activity and structural deficits in the fronto-limbic network but lacked predictive capability. On the other hand, He et al. (He et al., 2017) used a completely data-driven multimodal fusion method (mCCA + jICA) to jointly analyze functional connectivity and structural network alterations in MDD, achieving higher ML-based diagnostic accuracy in distinguishing patients from controls compared to single-modality ML models. Similarly, Guo et al. (Guo et al., 2024) combined individualized functional connectivity (IFC) from rs-fMRI and individualized structural connectivity (ISC) from diffusion tensor imaging (DTI). Although both studies employed advanced analytical methods and achieved promising diagnostic accuracies, they were limited by small sample sizes or the absence of cross-site validation, which restricted their generalizability to real-world applications.

### Key aims of this manuscript

Acknowledging these limitations, in the present study, we employed a whole-brain, data-driven multivariate fusion approach called transposed Independent Vector Analysis (tIVA) to investigate both joint and modality-unique function-structure network alterations in gray matter density (GMD) and ALFF in a relatively large cohort of MDD patients and HC. tIVA represents a significant advancement over previous fusion methods, such as mCCA+jICA, by addressing critical technical limitations inherent to these approaches (Lee et al., 2008). Unlike mCCA+jICA, which relies on maximizing linear correlations between modalities and is susceptible to random permutation ambiguity, tIVA jointly models cross-dataset dependencies through Source Component Vectors (SCVs) and leverages multivariate statistical dependencies, ensuring more robust and accurate source separation, particularly in multisite studies (Luo, 2023). Moreover, by leveraging structural and temporal covariation, tIVA captures independent brain networks, offering greater biological plausibility compared to atlas-based parcellations (Beckmann et al., 2005; Grecucci et al., 2023; Sui et al., 2023). Finally, this analysis was chosen for its capability to investigate alterations in brain network components that differentiate the two groups, using a noninvasive and task-free approach that removes performance-related confounds (Biswal et al., 1995).

Second, to enhance the generalizability of our findings to new cases and address the limitations of traditional group-level statistics inference methods, we employed a supervised machine learning (ML) classifier to extract a predictive model of MDD. A Random Forest approach was used to this aim. This approach was chosen not only for its robustness in handling high-dimensional data (Breiman, 2001; Hastie et al., 2009), but also for the possibility of identifying the most significant brain structural and functional components predictive of MDD pathology.

Building upon prior investigations (Qi et al., 2018; Gong et al., 2020; Gray et al., 2020; Opel et al., 2020), we expect to identify joint and modality-specific network components distinguishing MDD patients from HC. Precisely, we predict structural reductions in subcortical areas, including the insula and cingulate cortex, and functional reductions within cerebellar regions among MDD patients. Furthermore, we anticipate alterations in PFC activity, characterized by hyperactivity in the medial regions of the DMN and hypoactivity in the prefrontal lateral regions of the CEN. Notably, the primary benefit of conducting a joint analysis is elucidating abnormalities’ interdependence across modalities. Indeed, by leveraging joint components, this approach will reveal how deficits in one domain may influence or relate to those in another, offering a more integrated understanding of brain dysfunction in MDD.

## Materials and methods

### SRPBS Multi-disorder MRI Dataset

#### Dataset and Participants

Data collection and sharing for this project was provided by the DecNef Department at the Advanced Telecommunication Research Institute International, Kyoto, Japan and selected from the SRPBS Multi-disorder MRI Dataset (restricted version – https://bicr-resource.atr.jp/srpbs1600/). The dataset includes rs-fMRI and sMRI scans from a large sample of patients and controls who were clinically assessed by expert clinicians or teams according to DSM-IV-TR or DSM-5 criteria (Tanaka et al., 2021). To ensure a high standard of diagnostic reliability, the Mini-International Neuropsychiatric Interview (MINI) (Sheehan et al., 1998) was administered to participants in the Hiroshima, Showa and Tokyo sites. This procedure also ensured the exclusion of psychiatric disorders in the control group. All participants provided written informed consent for anonymous data sharing.

In this study, we included patients with major depression and health controls (HC). The two groups were balanced based on demographic characteristics (sex and age), and the data collection site used propensity score matching, resulting in an initial sample of 510 (MDD=255) participants. To ensure that only high-quality data was included in further analysis, a stringent quality control protocol excluded structural and functional brain images with artifacts, brain lesions, or abnormalities. This led to a final sample of 461 participants for the current study. Table 1 summarizes the demographic characteristics of participants of the included sample.

**Table 1.** Sample characteristics.

|  | HC (n=235) | MDD (n=226) | Statistics | p-Value |
| --- | --- | --- | --- | --- |
| Gender (male/female) | 116/119 | 116/110 | $\chi^2 = 0.11$ | 0.74 |
| Age (years) | 41.96 ± 13.44 | 42.39 ± 11.65 | $t = 0.372$ | 0.71 |
| Site | | | $\chi^2 = 4.12$ | 0.53 |
| HUH | 51 | 49 | $\chi^2 = 0.04$ | 0.84 |
| HRC | 19 | 15 | $\chi^2 = 0.47$ | 0.49 |
| HKH | 22 | 29 | $\chi^2 = 0.96$ | 0.33 |
| COI | 78 | 60 | $\chi^2 = 2.35$ | 0.13 |
| KUT | 13 | 14 | $\chi^2 = 0.04$ | 0.85 |
| UTO | 52 | 59 | $\chi^2 = 0.44$ | 0.51 |

**Abbreviations:** HUH, Hiroshima University Hospital; HRC, Hiroshima Rehabilitation Center; HKH, Hiroshima Kajikawa Hospital; COI, Hiroshima COI; KUT, Kyoto University TimTrio; UTO, University of Tokyo Hospital.

#### MRI acquisition

Structural and functional images were acquired using six 3T MRI scanners across multiple sites, according to the standardized SRPBS MRI acquisition guidelines (Tanaka et al., 2021). These protocols introduced recommendations for functional and structural image acquisition. For instance, rs-fMRI recommendations included but were not limited to full-brain coverage with particular attention to the cerebellum, minimized repetition time (TR), and a focus on prefrontal regions associated with psychiatric disorders. Similarly, structural imaging adhered to J-ADNI2 standards, emphasizing high-resolution isovoxel acquisition (1 × 1 × 1 mm) and full-brain coverage with sufficient margins. Moreover, during the scanning sessions, participants were instructed to remain relaxed, focus on a central crosshair displayed on a gray background, avoid engaging in specific thoughts or falling asleep, and minimize movement of their heads and trunks. Detailed imaging parameters for each site are provided in Table S1.

### MRI data processing

#### First-level preprocessing: sMRI and rs-fMRI

Multimodal brain imaging data were preprocessed following a rigorous quality control procedure resembling the Di and Biswal pipeline (Di and Biswal, 2023).

##### rs-fMRI preprocessing

Resting-state fMRI preprocessing steps were all performed in SPM12 (https://www.fil.ion.ucl.ac.uk/spm/) within MATLAB 2022b environment and included discarding the initial dummy scans, realigning to the first image, coregistering to the anatomical image, spatial normalization to MNI space, and spatial smoothing with an 8-mm FWHM isotropic Gaussian kernel. A stringent quality control protocol was employed, which included visual assessments for issues such as ghost artefacts, lesions, poor brain coverage, coregistration misalignments and normalization. Additionally, temporal domain issues, including head motion and other physiological noises, were evaluated. Specifically, framewise displacement (FD) was calculated based on rigid body transformation results to exclude participants with excessive head motion (FD > 1.5 mm or 1.5°). Overall, this procedure ended with the exclusion of 25 out of 510 participants.

Finally, the amplitude of low-frequency fluctuation (ALFF) maps was calculated for each participant from the preprocessed time series using the DPARSF-A (https://rfmri.org/DPARSF) (Yan and Zang, 2010). To do this, we regress out nuisance covariates, including mean signals from WM and CSF, as well as the Friston 24-parameter model (Friston et al., 1996), followed by bandpass filtering within a frequency range of 0.01–0.1 Hz to mitigate the impact of low-frequency drifts and high-frequency physiological noise (Yu-Feng et al., 2007).

##### sMRI preprocessing

Raw T1-weighted images were visually inspected across multiple slices in the x, y, and z planes to exclude scans with artifacts such as head motion, ghosting, structural abnormalities or lesions. Preprocessing was performed using CAT12 (http://dbm.neuro.uni-jena.de/cat/), an extension of SPM12 toolbox in the MATLAB 2022b environment (The MathWorks Inc., 2022). After the manual re-orientation of the anterior commissure as the origin, images were segmented into GM, white matter (WM), and cerebrospinal fluid (CSF). After segmentation, images were registered using Diffeomorphic Anatomical Registration Through Exponential Lie algebra (DARTEL) (Ashburner, 2007), normalized to Montreal Neurological Institute (MNI) standard space and smoothed with a 12-mm full-width at half-maximum (FWHM) isotropic Gaussian kernel. Given the multicenter design of the study, the CAT Quality Control framework was used to ensure consistent output preprocessing quality across scanners. As an outcome of this quality control procedure, an additional 24 participants were excluded from further analysis.

#### Transposed Independent Vector Analysis (tIVA)

A Transposed Independent Vector Analysis with a prior generalized Gaussian Distribution (tIVA-GGD) (Adali et al., 2015) was used to decompose individual whole-brain maps of GMD and ALFF into independent structural and functional networks. The tIVA-GGD, a data-driven multimodal blind source separation (BSS) technique, was chosen over other ICA algorithms or simple univariate methods for its capability of enhancing the identification and characterization of complex brain circuits that interact across different imaging modalities. Indeed, unlike standard ICA, IVA leverages statistical dependence across modalities while maintaining source independence within each modality (Lee et al., 2008; Laney et al., 2015; Luo, 2023). Furthermore, IVA incorporates higher-order statistics into the multiset canonical correlation analysis (mCCA) framework, offering a generalized approach that merges the strengths of both independent component analysis (ICA) and canonical correlation analysis (CCA) (Adali et al., 2015). The Minimum Description Length (MDL) method was used to estimate the number of components for each modality,(Li et al., 2007) yielding eight components. All analyses were conducted using the Fusion ICA Toolbox (FIT; version 2.0e; https://trendscenter.org/software/fit/).

The loading coefficient source matrix was reconstructed as a 3D image and scaled to a unit z-score for better visualization. Component 3, identified as noise in the GM component, was excluded from further analysis. Anatomical labels and stereotaxic coordinates were obtained from clusters exceeding a threshold of z = 3.5 by linking the IVA output images to the Talairach Daemon database (http://www.talairach.org/daemon.html). The results were then rendered using Surf Ice (https://www.nitrc.org/projects/surfice/) for better visualization. Refer to Figure 1 for a visual representation of the analysis workflow.

**Figure 1.**
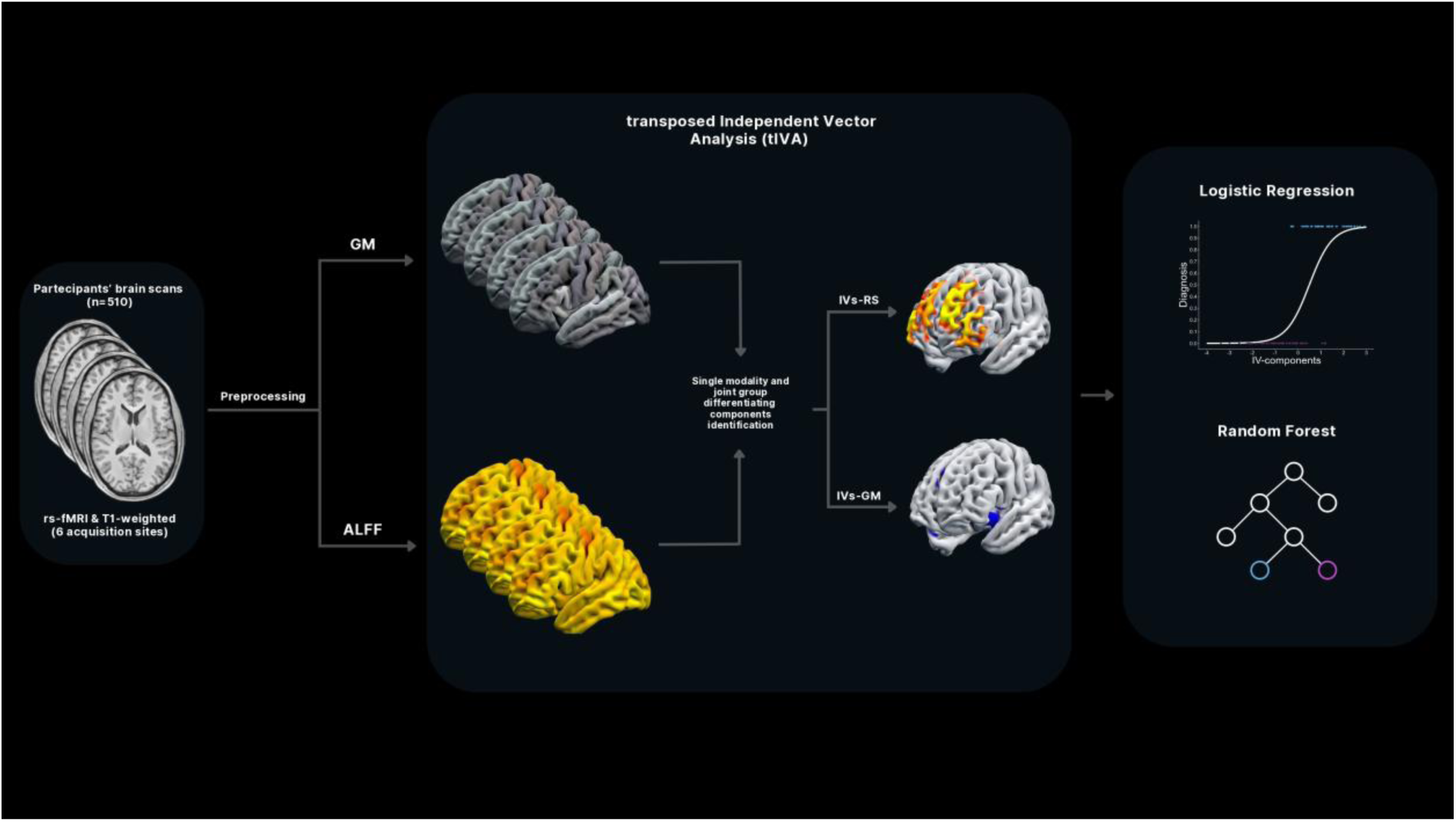
Multimodal Feature Extraction and Classification Pipeline. A data fusion unsupervised ML technique (tIVA) was used to decompose the brain into joint GM-ALFF independent networks after preprocessing structural and functional images. Afterwards, diagnosis labels were predicted using logistic regression and Random Forest classifier.

#### Logistic regression

A forward logistic regression model was implemented using JASP software (version 0.18.3.0; JASP Team, 2021) to identify significant transformed Independent Vectors (tIVs) predictors for distinguishing participants in the two groups. This method was chosen over backward and bidirectional stepwise regression due to its ability to incrementally build models based on statistical significance, thereby reducing the risk of overfitting. Indeed, forward logistic regression involves iteratively adding variables to an initially empty model based on their statistical significance, enabling robust and efficient feature selection.

#### Predictive Model

A supervised ML Random Forest classifier was employed to distinguish MDD patients from HC using tIVs as input features. RF was chosen over other supervised algorithms due to its robustness in handling overfitting and its superior performance with smaller sample sizes (Breiman, 2001; Hastie et al., 2009). Additionally, RFs implement feature importance ranking by measuring the contribution of each predictor to impurity reduction, thereby offering valuable interpretability in identifying brain regions as the most predictive of MDD. Functional and structural independent vectors (tIV-RS 1,2,4,6,7,8 and tIV-GM 1,2,4,6,7,8) were standardized separately for training and testing to prevent data leakage. The slight class imbalance in the dataset was effectively addressed using the Synthetic Minority Over-sampling Technique (SMOTE) (Chawla et al., 2002), which generates synthetic samples for the minority class through interpolation between existing data. This approach ensured that the RF model was trained on a balanced dataset, improving its generalisation ability across both classes. The implementation was from the imbalanced-learn Python package (Lemaître et al., 2017). Recursive Feature Elimination was applied to iteratively identify the most significant predictors by training the RF model and removing features that contributed the least to impurity reduction, a measure of data uncertainty. This process retained the most informative tIVs, enhancing model interpretability while ensuring robustness. The model’s performance was evaluated using metrics such as accuracy, precision, recall, specificity, F1 score, and the area under the Receiver Operating Characteristic (ROC) curve (AUC). An 80-20 train-test split was applied to ensure sufficient data for model training and evaluation. The Optuna framework optimised RF hyperparameters (Akiba et al., 2019), focusing on parameters like the number of trees, maximum depth, and minimum samples per split to enhance cross-validated accuracy. Additionally, nested cross-validation (5-fold inner for tuning and 5-fold and 10-fold outer for evaluation) was used to reduce partitioning bias and assess generalizability. Finally, permutation testing with 1,000 iterations further validated the model by ensuring the accuracy was not due to random label configurations (Ojala and Garriga, 2009).

### Data availability

The original data are available from the DecNef Department at the Advanced Telecommunication Research Institute International, Kyoto, Japan upon request.

## Results

### rs/s-MRI networks separating MDD from HC

The logistic regression model with the strongest explanatory value and significant improvement (*AIC* = 599.725, *BIC* = 620.392, *ΔΧ²* = 4.185, *P* = 0.041) achieved moderate accuracy (62.90%), exceeding the 50% random baseline in this balanced dataset. The model identified one joint component (tIV-GM7 and tIV-RS7) and two unique components (tIV-RS1 and tIV-RS2) as predictors.

The significant predictors were the joint-group differentiating components tIV-GM7 (*P* < 0.001) and tIV-RS7 (*P* < 0.001). Specifically, reduced subcortical GM concentration in tIV-GM7 regions, including the insula and cingulate cortex, significantly increases the probability of being diagnosed with MDD (*β* = –0.387, *SE* = 0.107, *Wald* = 13.174, *OR* = 0.679). Similarly, tIV-RS7, representing functional activity in the cerebellum, showed that reduced resting-state activity in these regions predicts a higher probability of MDD diagnosis (*β* = –0.371, *SE* = 0.106, *Wald* = 12.350, *OR* = 0.690).

The modality-unique components tIV-RS1 (*P* = 0.004) and tIV-RS2 (*P* = 0.037) were also significant. Notably, increased tIV-RS1 resting-state activity in regions primarily encompassing the dorsomedial prefrontal cortex (dmPFC) is associated with a higher probability of MDD diagnosis (*β* = 0.261, *SE* = 0.100, *Wald* = 6.761, *OR* = 1.298). Conversely, decreased tIV-RS2 resting-state activity in regions predominantly involving the dorsolateral prefrontal cortex (dlPFC) heightened the likelihood of MDD diagnosis (*β* = –0.202, *SE* = 0.100, *Wald* = 4.618, *OR* = 0.817). Table 2 and 3 provides the complete Talairach coordinates for all significant tIVs, while Figures 2 and 3 visually represent the identified significant components.

**Figure 2.**
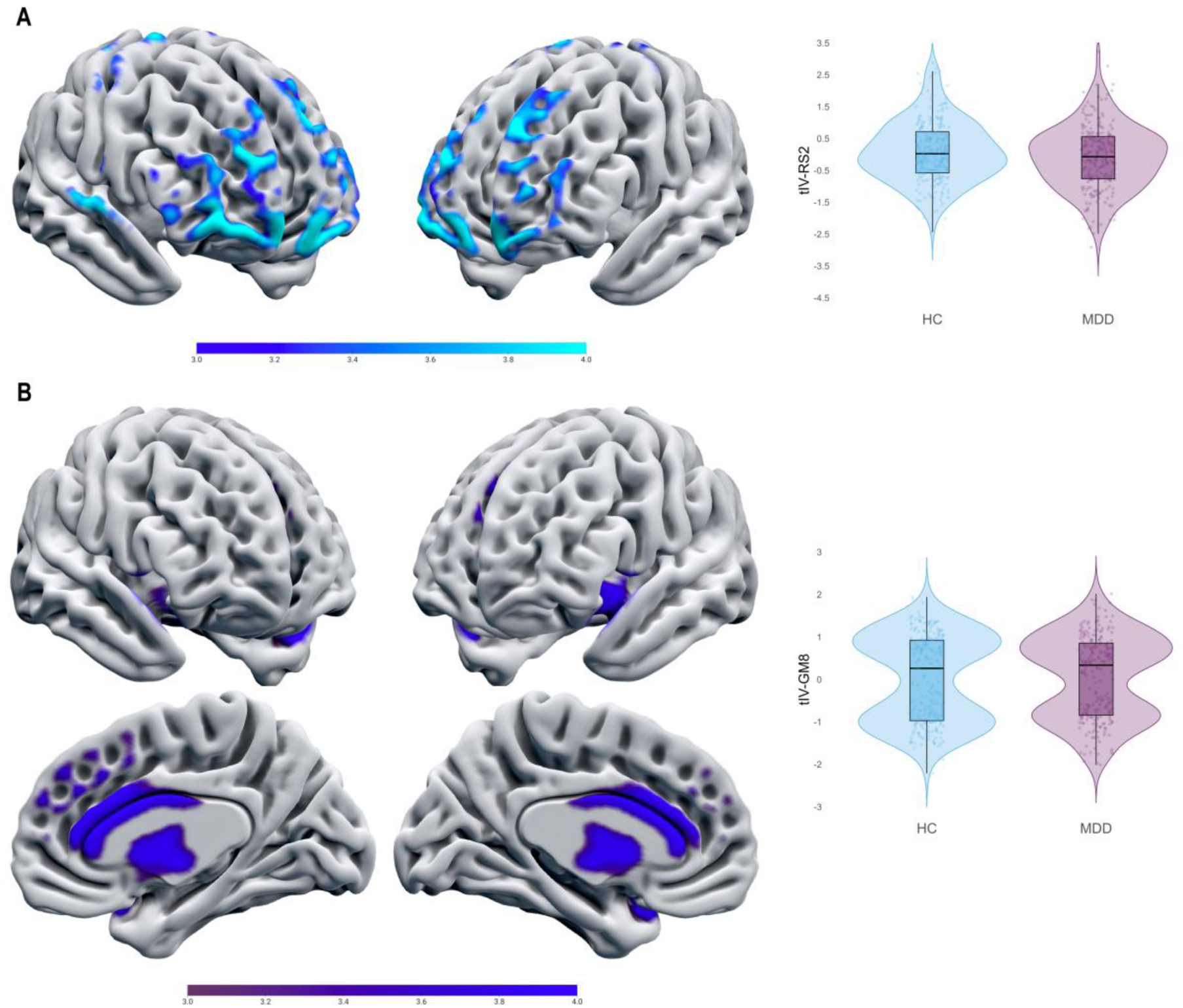
Joint functional and structural tIV-7 alterations in MDD. **(A)** Visual representation of tIV-RS7, revealing reduced ALFF (blue-light blue scale) in the cerebellar tonsil and inferior semi-lunar lobule among MDD patients. **(B)** Visual representation of tIV-GM7, indicating reduced GM density (violet-blue scale) in regions of the SN among MDD patients. The violin plots on the right compare activation and GM concentration differences between controls and depressed patients.

**Figure 3.**
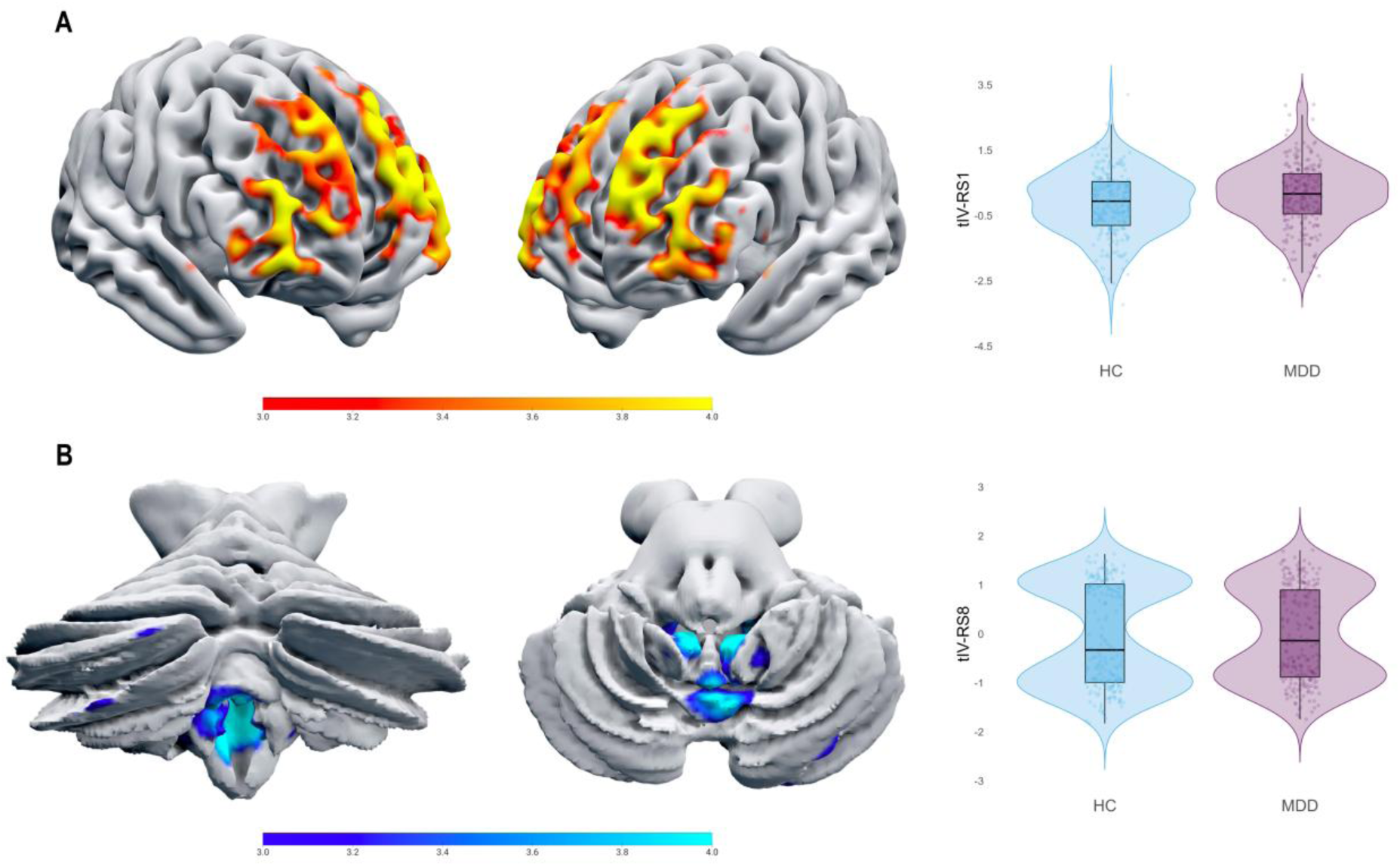
Single-modality group-differentiating components tIV-RS1 and tIV-RS2. **(A)** Visual representation of tIV-RS1, highlighting hyperactivation (red-yellow scale) within the dmPFC among MDD patients. **(B)** Visual representation of tIV-RS2, indicating hypoactivation (blue-light blue scale) in the dlPFC among MDD patients. The violin plots on the right compare activation level differences between controls and depressed patients.

**Table 2.** Talairach coordinates of joint-modality brain circuit associated with Major Depressive Disorder.

| Area | Brodman Area | Volume (cc) | Random Effects: Max Value (x, y, z) |
| --- | --- | --- | --- |
| **tIV-RS7** |  |  |  |
| Cerebellar Tonsil | * | 0.8/0.4 | 6.0 (-3, -50, -42)/4.7 (4, -52, -42) |
| Lingual Gyrus | 17, 18 | 1.5/1.9 | 5.5 (-1, -85, -10)/5.7 (3, -85, -10) |
| Inferior Occipital Gyrus | 17, 18 | 0.3/0.6 | 4.3 (-28, -85, -13)/5.6 (13, -92, -8) |
| Precuneus | 7, 19, 31 | 0.8/1.2 | 4.9 (-6, -78, 45)/5.4 (1, -74, 40) |
| Fusiform Gyrus | 18, 19 | 0.6/0.4 | 4.6 (-19, -90, -14)/5.4 (22, -91, -12) |
| Cuneus | 7, 17, 18, 19, 23, 30 | 3.1/2.2 | 5.3 (-1, -75, 37)/4.7 (1, -78, 20) |
| Inferior Semi-Lunar Lobule | * | 0.4/0.1 | 4.8 (-1, -65, -37)/4.4 (1, -62, -37) |
| Uvula of Vermis | * | 0.2/0.1 | 4.4 (-1, -64, -33)/4.1 (1, -61, -34) |
| Medial Frontal Gyrus | 11 | 0.1/0.0 | 4.1 (-4, 60, -18)/-999.0 (0, 0, 0) |
| Declive | * | 0.2/0.0 | 4.0 (-37, -82, -15)/-999.0 (0, 0, 0) |
| Posterior Cingulate | 30 | 0.1/0.3 | 3.5 (-7, -68, 10)/3.9 (9, -66, 13) |
| Nodule | * | 0.1/0.0 | 3.9 (-7, -50, -30)/-999.0 (0, 0, 0) |
| Precentral Gyrus | 4, 6 | 0.0/0.3 | -999.0 (0, 0, 0)/3.9 (31, -19, 66) |
| Tuber | * | 0.1/0.2 | 3.8 (-34, -86, -27)/3.7 (33, -86, -27) |
| Middle Occipital Gyrus | 18 | 0.1/0.1 | 3.5 (-30, -90, 2)/3.8 (25, -85, -8) |
| Uvula | * | 0.2/0.1 | 3.8 (-6, -64, -33)/3.6 (4, -64, -33) |
| Pyramis | * | 0.1/0.0 | 3.6 (-39, -76, -34)/-999.0 (0, 0, 0) |
| **tIV-GM7** |  |  |  |
| Inferior Frontal Gyrus | 13, 47 | 2.4/2.4 | 7.0 (-40, 15, -13)/6.7 (42, 15, -11) |
| Superior Temporal Gyrus | 22, 38 | 4.0/3.0 | 6.7 (-45, 11, -8)/6.8 (45, 13, -8) |
| Insula | 13 | 1.7/1.2 | 6.4 (-45, 8, -5)/5.4 (43, 7, -4) |
| Sub-Gyral | 13, 21 | 0.2/0.1 | 6.2 (-42, 8, -8)/4.6 (43, 1, -9) |
| Extra-Nuclear | 13 | 1.1/1.4 | 5.9 (-39, 11, -9)/5.8 (43, 7, -9) |
| * | 10 | 0.4/0.4 | 4.8 (0, 44, 30)/4.7 (1, 28, 11) |
| Medial Frontal Gyrus | 6, 8, 9, 10 | 1.2/1.7 | 5.4 (0, 42, 20)/4.9 (3, 42, 17) |
| Anterior Cingulate | 24, 25, 32, 33 | 2.1/1.9 | 5.4 (0, 36, 20)/5.0 (3, 41, 12) |
| Caudate | * | 1.2/1.0 | 5.1 (-7, 8, 8)/4.7 (9, 9, 9) |
| Third Ventricle | * | 0.0/0.5 | -999.0 (0, 0, 0)/4.5 (0, -9, -1) |
| Cingulate Gyrus | 24, 32 | 1.0/1.0 | 4.2 (0, 13, 30)/4.4 (1, 21, 39) |
| Lateral Ventricle | * | 0.5/0.4 | 4.3 (-3, 10, 2)/4.3 (3, 7, 2) |
| Superior Frontal Gyrus | 8, 9 | 0.6/0.3 | 4.2 (0, 20, 49)/4.1 (3, 55, 22) |
| Uncus | 28 | 0.1/0.1 | 3.6 (-27, 6, -20)/3.8 (28, 9, -19) |
| Thalamus | * | 0.2/0.1 | 3.7 (-4, -3, 6)/3.5 (6, -10, 13) |

**Note:** Voxels with z-values greater than 3.5 were matched with the Talairach Daemon database to obtain anatomical labels and converted into MNI space. For each hemisphere, the maximum z-value and MNI coordinate are provided. The volume of voxels in each area is given in cubic centimetres (cc) (\*) = area not recognized by standard Brodmann Area atlas.

**Table 3.**
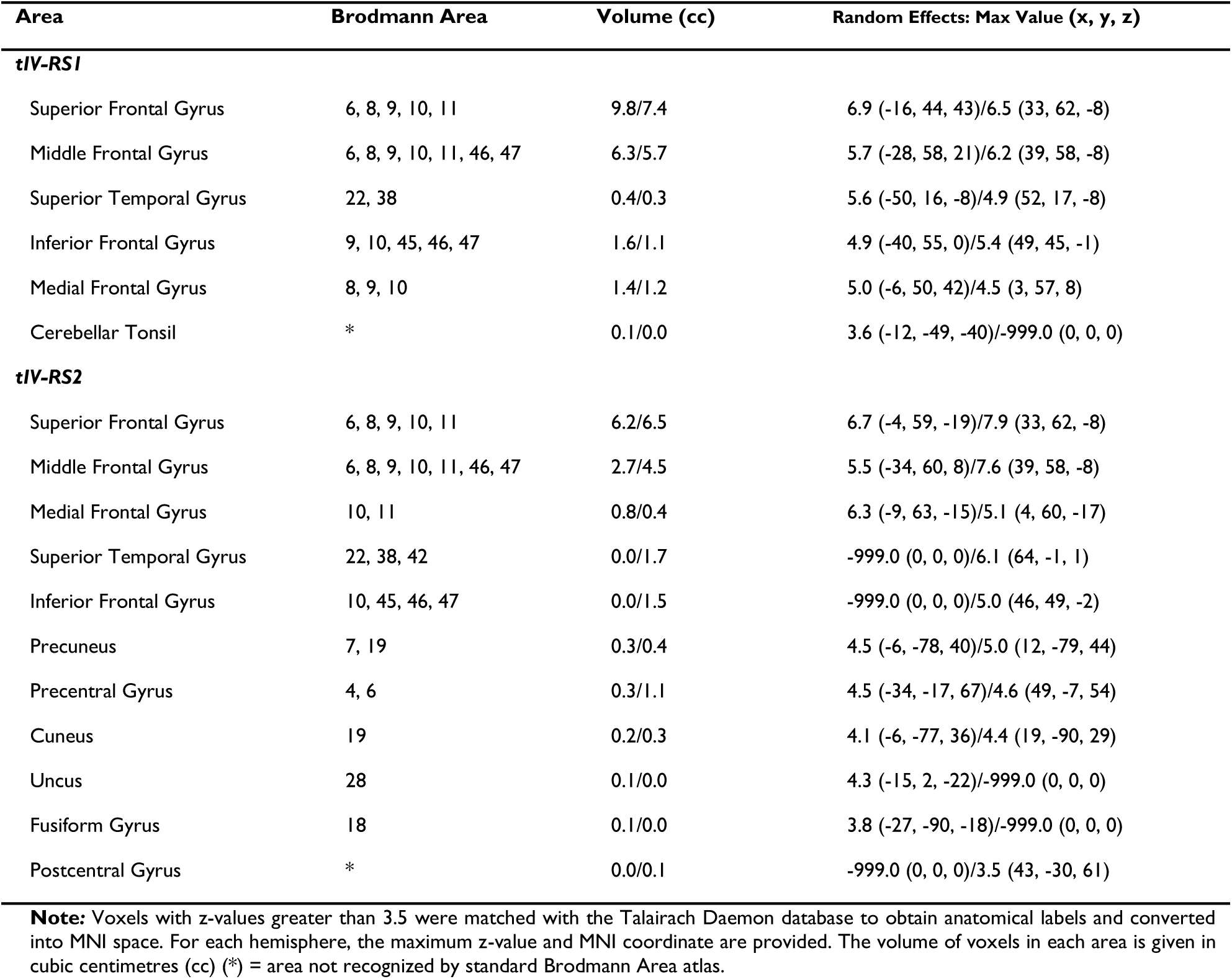
Talairach coordinates of single modality brain circuit associated with Major Depressive Disorder.

### RF predictive model

The RF classifier effectively distinguished participants in the two groups, with tIV-GM7 and tIV-RS7 emerging as the most important predictors, thus confirming the regression results, followed by tIV-RS8, tIV-RS2, tIV-GM8, and tIV-GM4 (Figure 4A), partially aligning and extending regression results. Feature importance analysis (Table S2), complete Talairach coordinates (Table S3), and visual representation of additional components (Figure S1, Figure S2) are provided in the supplementary materials. The model achieved a training accuracy of 81.08% (HC Precision/Recall: 80.10% / 82.70%; MDD Precision/Recall: 82.12% / 79.46%) and a test accuracy of 69.89% (HC Precision/Recall: 75.00% / 66.00%; MDD Precision/Recall: 65.31% / 74.42%). Figure 4B presents the confusion matrix for these results. Cross-validation further validated the model’s performance (5-fold: 69.94%; 10-fold: 68.00%) as shown in Figure 4C. Permutation tests confirmed the statistical significance of these accuracies (*P* < 0.001), while ROC curve analysis (Figure 4D) supported good class separation.

**Figure 4.**
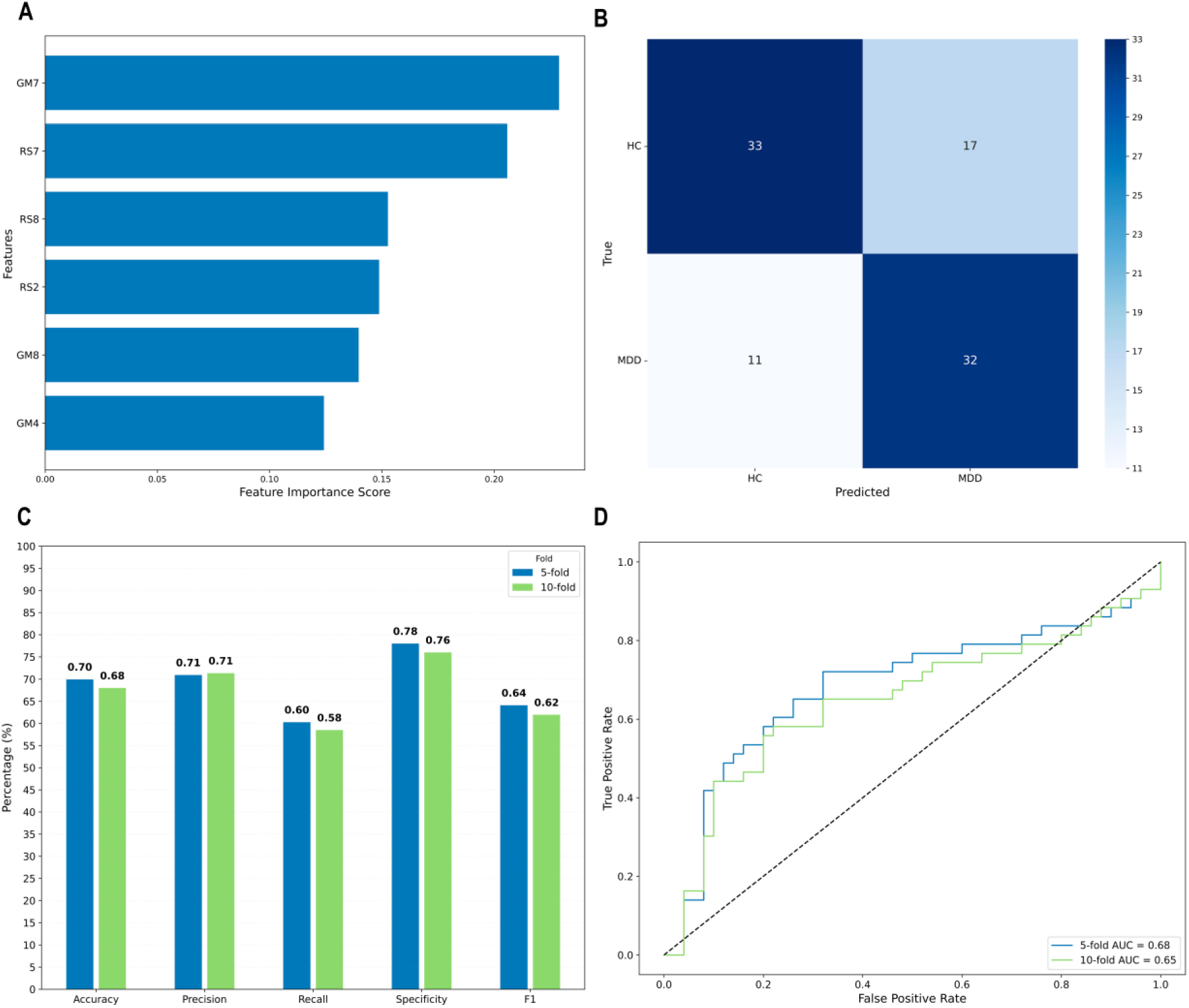
Visualization of Random Forest model performance. **(A)** Feature importance ranking, identifying the most influential features (GM7, RS7, RS6, RS2, GM6, and GM4) for classification. **(B)** Confusion matrix depicting true and predicted classifications for healthy controls (HC) and Major Depressive Disorder (MDD) patients in the test set. **(C)** Performance metrics (Accuracy, Precision, Recall, Specificity, and F1 Score) evaluated using 5-fold and 10-fold cross-validation, demonstrating consistent results across both validation strategies. **(D)** ROC curves comparing model performance under 5-fold and 10-fold cross-validation. The x-axis represents the false positive rate, and the y-axis represents the true positive rate. The diagonal dashed line represents the performance of a random classifier.

## Discussion

This study employs a data-driven tIVA multimodal fusion approach to examine interactions between structural (GMD) and functional (ALFF) alterations, offering deeper insights into the neurobiological mechanisms of MDD. Data from 461 participants, matched for age, sex, and MRI acquisition site, were used. The analysis identified key distinctive features of MDD across cerebellar regions, the SN and two modality-unique components. Finally, a Random Forest classifier identified tIV-RS2, tIV-RS7, tIV-RS8, tIV-GM4, tIV-GM7, and tIV-GM8 as the most significant predictors of MDD, achieving a test accuracy of 69.89%. Among these, tIV-GM7 (SN) was the most critical feature, followed by tIV-RS7 (cerebellum) and other joint and modality-unique components.

### Joint components: Cerebellum and SN

Our results showed a key joint tIV-7 component differentiating MDD from HC, revealing underlying associations between functional disruptions in cerebellar regions (tIV-RS7) and structural deficits in the SN (tIV-GM7). Notably, MDD participants exhibited both reduced GM concentration in the SN and reduced cerebellar activity.

Traditionally recognized for its role in motor control (Hariri, 2019), the cerebellum has been investigated for its involvement in neuropsychiatric conditions, such as the cerebellar cognitive and affective syndrome (Schmahmann et al., 2007). Indeed, a growing body of evidence has elucidated how this region plays a significant role in a variety of non-motor functions, including mood regulation, emotion processing, and social cognition (Stoodley and Schmahmann, 2010; Adamaszek et al., 2017; Depping et al., 2018; Schmahmann, 2019; Van Overwalle et al., 2020; Zanella et al., 2022). This broader role of the cerebellum is corroborated by connectivity analyses, revealing its functional interaction with all major brain networks, including the SN (Seeley et al., 2007; Van Overwalle et al., 2015).

In MDD, consistent evidence points to structural (Balcioglu and Kose, 2018) and functional (Alalade et al., 2011; Guo et al., 2013; Depping et al., 2018) disruptions in cerebellar components. Although the exact role of cerebellar dysfunctions in depressive symptoms remains debated (Phillips et al., 2015), they appear to strongly impact mood-related information processing, leading to an intensification of negative mood symptomatology (Clausi et al., 2019). These considerations make the cerebellum a potential target for therapeutic intervention in depression (Depping et al., 2017; Miterko et al., 2019). On the other hand, structural results are supported by findings from multi-centre studies emphasizing the SN’s pivotal role in depressive psychopathology (Ancelin et al., 2019). On top of that, the structural abnormalities observed correspond to gene expression changes associated with MDD, as Qi et al. (Qi et al., 2018) elucidated. Using a multimodal neuroimaging and transcriptomics fusion model (MCCAR+jICA), researchers identified microglia and neuronal transcriptional changes as key contributors to structural and functional integrity issues in fronto-limbic network regions, including the dlPFC and ACC.

These relevant works provide compelling evidence-based framing, providing additional validity to our data-informed insights about the cerebellum and SN patterns.

### Modality unique components: DMN and CEN

Two modality-unique components — tIV-RS1 and tIV-RS2 — significantly contributed to differentiating MDD patients from HC. tIV-RS1 primarily includes brain circuits within the dorsomedial prefrontal regions of the DMN, while tIV-RS2 is more associated with dorsolateral regions of the CEN. Interestingly, opposing activation patterns were observed, with MDD patients showing significantly increased activity in tIV-RS1 and decreased activity in tIV-RS2.

The findings for the dmPFC component tIV-RS1 are consistent with previous studies that underscored the DMN’s critical involvement in MDD psychopathology, exhibiting enhanced functional connectivity and activity (Hamilton et al., 2015; Kaiser et al., 2015; Scheepens et al., 2020). The DMN is a task-negative network engaged in introspective processes such as mind-wandering(Mason et al., 2007; Andrews-Hanna, 2012) and self-referential thought (Raichle et al., 2001; Whitfield-Gabrieli et al., 2011; Davey et al., 2016). It also contributes to emotional regulation (Pan et al., 2018), social cognition (Schilbach et al., 2008), and episodic memory (Spreng et al., 2009). By integrating these functions, the DMN supports the formation of a unified ‘internal narrative,’ allowing individuals to reflect on and make sense of their personal experiences (Menon, 2023).

In people with MDD, the internal narrative often becomes overly negative and persistent, contributing to the development and reinforcement of rumination. This maladaptive psychological process has been linked to functional hyperactivation in the dmPFC subsystem of the DMN (Hamilton et al., 2011; Zhou et al., 2020) a finding corroborated by our results. Furthermore, alterations in the dmPFC appear to be related to impairments in mentalizing and metacognition in MDD (Nestor et al., 2022), disrupting the ability to integrate social information with personal beliefs and emotions, as well as to understand others’ mental states (Lieberman, 2007; Van Overwalle, 2009; Lombardo et al., 2010). This heterogeneous symptomatology of MDD has been explained by Scalabrini et al. (Scalabrini et al., 2020) as resulting from global connectivity disruptions, where hyperactivity of the DMN destabilizes non-DMN networks and exacerbates depressive symptoms.

In contrast to DMN hyperactivity, our findings indicate a dlPFC hypoactivation in the CEN at the level of the tIV-RS2. The CEN plays a crucial role in task-positive functions such as cognitive control, goal-directed behaviour (Miller and Cohen, 2001; Fuster, 2006; Dalley et al., 2011) working memory,(D’Esposito and Postle, 2015), and spatial attention (Corbetta and Shulman, 2002). Disruptions in this network have been extensively reported across various psychiatric conditions, such as depression, Alzheimer’s disease, and schizophrenia (Brewin et al., 2010; Woodward et al., 2011; Zhao et al., 2019). Our findings of dlPFC hypoactivation are consistent with existing literature linking this region to deficits in attention, emotional processing, and self-referential processing in MDD (Lemogne et al., 2010; Sánchez-Navarro et al., 2014). Notably, different evidence suggests that dlPFC hypoactivity may significantly influence attentional and emotional biases, particularly by promoting an excessive focus on internally directed thoughts like rumination (Northoff, 2007; Plewnia et al., 2015). This reduced dlPFC is closely associated with disrupted connectivity with the subgenual anterior cingulate cortex (sgACC), a key region for emotional regulation. Given the disrupted dlPFC-sgACC connectivity, transcranial magnetic stimulation (TMS) targeting the dlPFC has shown promise in alleviating MDD symptomatology, particularly in cases resistant to other therapies (Padberg and George, 2009; Fox et al., 2012; Weigand et al., 2018).

Overall, the hypoactivity of the CEN and hyperactivity of the DMN in MDD reflect a disrupted balance between intra– and inter-network dynamics, leading to an unbalance between externally directed cognitive control and internally focused cognition (Sporns, 2013; Shine and Poldrack, 2018; Menon and D’Esposito, 2022). According to Menon’s triple network model of psychopathology, the SN is critical for switching between the DMN and CEN in response to external demands (Menon, 2011; Menon and D’Esposito, 2022). However, in MDD, aberrant integration and segregation processes in SN hubs such as the dorsal anterior cingulate cortex (dACC) and anterior insula (AI) impair this switching process, further exacerbating cognitive and emotional dysfunctions (Dosenbach et al., 2008; Seeley, 2019; Krönke et al., 2020; Caria and Grecucci, 2023). This breakdown can be further understood through the co-occurrence of structural and functional abnormalities identified in this study. Structural deficits in the SN impair its ability to regulate dynamic switching between the DMN and CEN, leading to dysfunction in cerebellar regions critical for mood and cognitive regulation. In turn, reduced cerebellar activity exacerbates maladaptive feedback to cortical networks, intensifying the emotional and cognitive impairments central to MDD. These joint modality findings underscore the interconnected roles in pathological emotional regulation and cognitive processes central to MDD and the importance of integrating structural and functional data to elucidate the mechanisms underlying MDD psychopathology.

In conclusion, the single-modality rs-fMRI results discussed are primarily consistent with the meta-analysis by Gong et al. which reported increased mPFC activity and reduced cerebellum activity among MDD patients (Gong et al., 2020). The structural results aligned with insights from single-modality and multimodal meta-analyses, which have identified overlapping structural abnormalities, such as reduced GMD in the SN (tIV-GM7), especially in the insula and anterior cingulate cortex (Gray et al., 2020; Opel et al., 2020).

### Interpretation of other insights

The RF model corroborates tIV-GM7, tIV-RS7, and tIV-RS2 as critical brain components for distinguishing the two groups, achieving a test accuracy of 69.89%. In addition to these primary features, other components — tIV-GM4, tIV-RS8, and tIV-GM8 — were also identified as significant contributors to the RF model. tIV-GM4 involves regions such as the precuneus, cuneus, and thalamus, while the joint components, tIV-GM8 and tIV-RS8, contributed significantly to the classification model. This joint component reflects functional disruptions in posterior cortical regions (tIV-RS8) alongside structural abnormalities in the thalamus and cerebellar lingual gyrus (tIV-GM8). While these components did not emerge as significant predictors in the logistic regression analysis, their inclusion in the RF model underscores their relevance to the broader neural abnormalities underlying MDD.

Considering these ML results, the multimodal fusion approach used here demonstrates the potential advantage of integrating both structural and functional features for a more nuanced and more accurate MDD prediction. Indeed, comparing our results to existing classification rs-fMRI studies, our ML test performance is aligned with or surpasses studies using similar sample sizes (Bondi et al., 2023). Furthermore, our approach remains competitive against large-scale efforts, such as the ENIGMA consortium study by Belov et al., which reported balanced test accuracies between 52% and 63% using only structural data (Belov et al., 2024).

However, it is important to note that smaller-scale studies often report inflated accuracies — up to 95% in some cases — due to methodological flaws like improper cross-validation, data leakage, and overfitting (Flint et al., 2021; Bondi et al., 2023). These pitfalls underscore the critical role of robust validation and large independent test sets. Additionally, Belov et al. (Belov et al., 2024) highlighted the limitations of relying solely on sMRI in multi-site studies, which can result in poor classification performance.

## Limitation and Future Direction

Notwithstanding the above valuable insights, this study has limitations. First, the relatively homogeneous Japanese cohort may limit the generalizability of the findings to other ethnic groups. While, to the best of our knowledge, this is the first multimodal fusion study of this type (ALFF+GMD) conducted on this population, the homogeneity of the sample may limit the generalizability of the results to other ethnic groups. Specifically, population differences in MDD have been consistently documented, including variations in prevalence rates,(Ferrari et al., 2013) expression of psychosomatic symptoms (Ryder et al., 2008), and distinct risk genes (Bigdeli et al., 2017). As Chen et al. emphasized, a crucial future direction for large-scale projects will be to address cultural and ethnic disparities by incorporating cross-cultural samples through international collaborations (Chen et al., 2022b). Second, while multisite studies may introduce site-specific noise and scanner-related artifacts, they also provide an opportunity to train ML models under conditions resembling more closely real-world scenarios, enhancing results’ generalizability (Bento et al., 2022; Chen et al., 2022a). Moreover, to mitigate scanner batch effects, we employed tIVA, a highly robust and effective method for multisite studies, proven to extract reliable components under these conditions (Luo, 2023). The stable performance of our ML models, validated through cross-validation and permutation tests, further confirms the robustness of these findings to inter-site variability. Third, although the sample was balanced for age, sex, and acquisition site, other variables such as socioeconomic status, lifestyle, and medication treatment were not controlled due to the absence of this information in the original dataset. While the influence of these variables on brain structure and functional biomarkers remains debated (Enneking et al., 2020; Mohammadi et al., 2023; Zhao et al., 2023), they are increasingly recognized as critical factors in the development of ML models for clinical applications (Williams and Whitfield Gabrieli, 2024). Therefore, future multisite efforts should prioritize collecting such data to enhance models’ robustness and applicability. Finally, transdiagnostic fusion studies, such as Qi et al., which identify shared brain networks across schizophrenia, MDD, and other conditions, may be preferred to provide valuable insights into overlapping and different neural mechanisms in psychiatric conditions (Qi et al., 2020).

## Conclusion

The current study demonstrates a critical interaction between functional hypoactivation in cerebellar regions and structural deficits in SN regions in MDD, highlighting a central role of these multimodal alterations in the pathological emotional regulation and cognitive impairments characterizing this disorder. Furthermore, leveraging structural and functional independent vector components, a Random Forest classifier achieved nearly 70% accuracy in distinguishing MDD patients from HC. These findings highlight the potential of ML combined with data-driven fusion techniques for identifying neuroimaging biomarkers and advance precision diagnostics and treatment in clinical psychiatry.

## Supporting information

Supplemental Images and Tables

## Acknowledgments

Data collection and sharing for this project were provided by the DecNef Department at the Advanced Telecommunication Research Institute International (ATR), Kyoto, Japan, as part of the SRPBS Multi-disorder MRI Dataset (restricted version). The dataset is available at https://bicr-resource.atr.jp/srpbs1600/.

