## Supplemental Images and Tables for "The dark side of the mood: structural and functional fronto-insular and cerebellar alterations classify major depression"

### Supplemental Materials

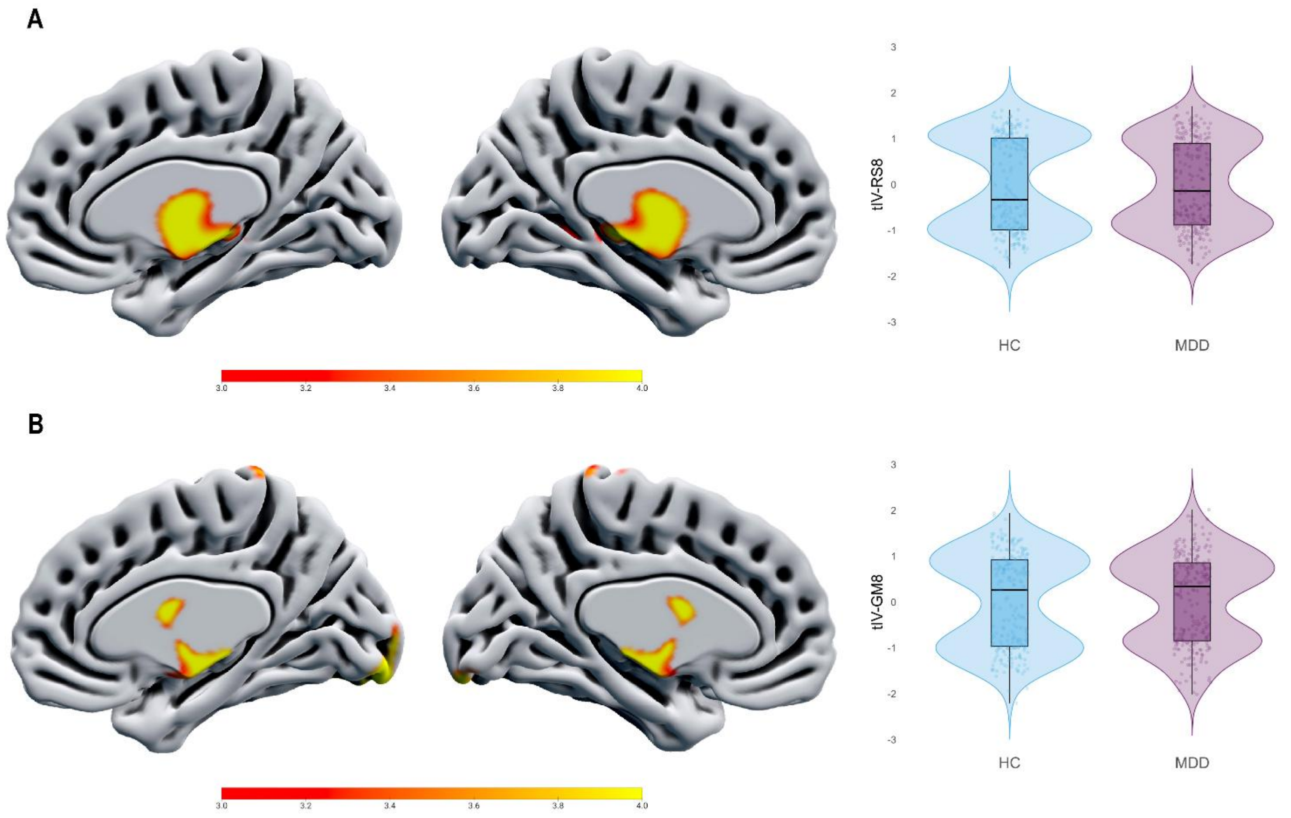


**Figure S1. Joint functional and structural tIV-8 alterations in MDD.** **(A)** Visual representation of tIV-RS8, revealing increased ALFF (red-yellow scale) in salience network regions among MDD patients. **(B)** Visual representation of tIV-GM8, indicating increased GM density (red-yellow scale) in the thalamus among MDD patients. The violin plots on the right compare activation and GM concentration differences between controls and depressed patients.

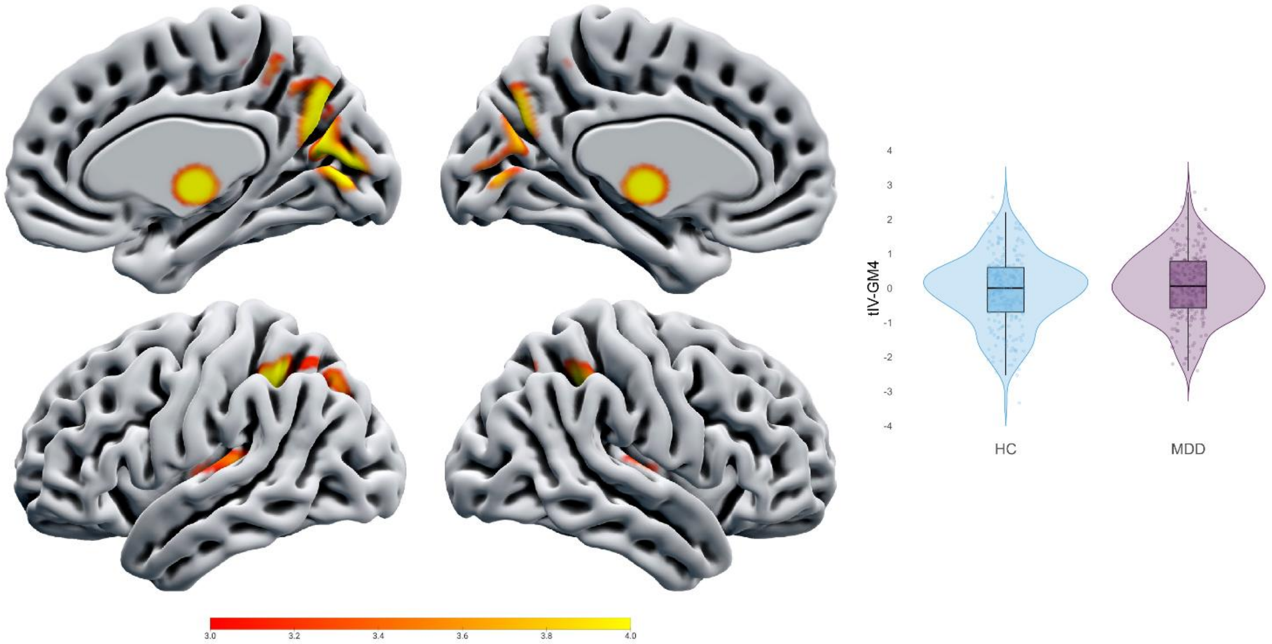


**Figure S2. Single-modality group-differentiating component tIV-GM4. (A)** Visual representation of tIV-GM4, depicting increased GM density (red-yellow scale) within the parietal cortex, including the precuneus and inferior parietal lobule among MDD patients. The violin plots on the right compare GM concentration differences between controls and depressed patients.

**Table S1. Image acquisition parameters for rs-MRI and s-MRI.**

| Site | HUH | HRC | HKH | COI | KUT | UTO |
|---|---|---|---|---|---|---|
| MRI scanner | GE Signa HDxt | GE Signa HDxt | Siemens Spectra | Siemens Verio.Dot | Siemens TimTrio | GE MR750w |
| ***rs-fMRI parameters*** | | | | | | |
| Magnetic field strength | 3.0T | 3.0T | 3.0T | 3.0T | 3.0T | 3.0T |
| No.of channels per coil | 8 | 8 | 12 | 12 | 32 | 24 |
| FoV, mm | 212 | 256 | 192 | 212 | 212 | 256 |
| Matrix | 64 × 64 | 64 × 64 | 64 × 64 | 64 × 64 | 64 × 64 | 64 × 64 |
| No. of slices | 32 | 32 | 38 | 40 | 40 | 40 |
| No. of volumes | 143 | 143 | 107 | 240 | 240 | 240 |
| In-plane resolution | 3.3 × 3.3 | 4.0 × 4.0 | 3.0 × 3.0 | 3.3 × 3.3 | 3.3125 × 3.3125 | 3.3 × 3.3 |
| Slice thickness | 3.2 | 4 | 3 | 3.2 | 3.2 | 3.2 |
| Slice gap | 0.8 | 0 | 0 | 0.8 | 0.8 | 0.8 |
| TR, ms | 2,500 | 2,000 | 2,700 | 2,500 | 2.500 | 2,500 |
| TE, ms | 30 | 27 | 31 | 30 | 30 | 30 |
| Total scan time, min:s | 10:17 | 04:46 | 04:49 | 10:00 | 10:00 | 10:00 |
| Flip angle, deg | 80 | 90 | 90 | 90 | 80 | 80 |
| Slice acquisition order | Ascending | Ascending | Ascending | Ascending | Ascending | Ascending |
| Phase encoding | PA | PA | AP | AP | PA | PA |
| Eyes closed/fixated. | Fixated | Fixated | Fixated | Fixated | Fixated | Fixated |
| ***sMRI parameters*** | | | | | | |
| FoV | 256 | 256 | 256 | 256 | 225 × 240 | 240 |
| Matrix | 256 × 256 | 256 × 256 | 256 × 256 | 256 × 256 | 240 × 256 | 256 × 256 |
| Voxel size | 1 × 1 × 1 | 1 × 1 × 1 | 1 × 1 × 1 | 1 × 1 × 1 | 0.9375× 0.9375×1.0 | 1 × 1 × 1.2 |
| TR | 6812 | 6812 | 1900 | 2300 | 2000 | 7.7 |
| TE | 1896 | 1896 | 2.38 | 2.98 | 3.4 | 3.1 |
| TI | 450 | 450 | 900 | 900 | 990 | 900 |
| Flip angle | 20 | 20 | 10 | 9 | 8 | 11 |

**Abbreviations:** HUH, Hiroshima University Hospital; HRC, Hiroshima Rehabilitation Center; HKH, Hiroshima Kajikawa Hospital; COI, Hiroshima COI; KUT, Kyoto University TimTrio; UTO, University of Tokyo Hospital; FoV: Field of view; TR, repetition time; AP, anterior to posterior; PA, posterior to anterior; TE, echo time; TI, inversion time; HUH, Hiroshima University Hospital; HRC, Hiroshima Rehabilitation Center; HKH, Hiroshima Kajikawa Hospital; COI, Hiroshima COI; KUT, Kyoto University TimTrio; UTO, University of Tokyo Hospital.

**Table S2. Feature importance for brain components of Random Forest classification**

| Rank | Feature | Mean Decrease in Gini Impurity |
|---|---|---|
| 1 | GM7 | 0.2289 |
| 2 | RS7 | 0.2059 |
| 3 | RS8 | 0.1527 |
| 4 | RS2 | 0.1487 |
| 5 | GM8 | 0.1396 |
| 6 | GM4 | 0.1241 |

**Table S3. Talairach coordinates of the brain circuit associated with MDD**

| Area | Brodmann Area | Volume (cc) | Random Effects: Max Value (x, y, z) |
|---|---|---|---|
| ***tIV-GM4*** | | | |
| Third Ventricle | * | 0.0/0.4 | -999.0 (0, 0, 0)/5.3 (0, -15, -1) |
| Precuneus | 7, 19, 23, 31 | 2.9/3.3 | 4.7 (-1, -66, 25)/5.1 (1, -67, 36) |
| Cuneus | 7, 17, 18, 23 | 0.6/1.4 | 4.8 (-1, -68, 32)/5.0 (3, -77, 12) |
| Inferior Parietal Lobule | 40 | 2.3/0.8 | 4.9 (-34, -43, 45)/4.3 (36, -42, 44) |
| Sub-Gyral | * | 1.2/0.5 | 4.8 (-34, -40, 42)/3.9 (33, -42, 41) |
| Thalamus | * | 0.3/0.3 | 4.4 (-3, -14, 2)/4.6 (3, -14, 2) |
| Cingulate Gyrus | 31 | 0.3/0.5 | 4.1 (-1, -46, 42)/4.6 (1, -46, 40) |
| Extra-Nuclear | * | 0.2/0.3 | 4.3 (-3, -18, 0)/4.6 (3, -18, 0) |
| Insula | 13, 22 | 0.9/1.0 | 4.4 (-45, -6, 0)/4.3 (45, -9, 3) |
| Lingual Gyrus | 18 | 0.1/0.1 | 3.7 (-1, -83, 4)/4.3 (1, -77, 5) |
| Posterior Cingulate | 30, 31 | 0.1/0.4 | 3.8 (-1, -60, 25)/4.2 (3, -69, 15) |
| Superior Parietal Lobule | 7 | 0.5/0.1 | 4.2 (-33, -49, 48)/3.5 (28, -63, 49) |
| Superior Temporal Gyrus | 13, 22, 38 | 0.4/0.5 | 4.0 (-48, -9, 2)/3.9 (45, 1, -4) |
| Paracentral Lobule | 5, 31 | 0.3/0.2 | 4.0 (0, -40, 50)/3.9 (1, -34, 50) |
| Transverse Temporal Gyrus | 41 | 0.3/0.0 | 4.0 (-39, -23, 11)/-999.0 (0, 0, 0) |
| Precentral Gyrus | * | 0.1/0.0 | 3.8 (-46, -11, 6)/-999.0 (0, 0, 0) |
| ***tIV-GM8*** | | | |
| * | * | 0.1/0.2 | 4.3 (-3, -6, -3)/4.9 (1, -1, 4) |
| Thalamus | * | 2.2/1.7 | 5.3 (-4, -7, 9)/5.7 (3, -6, 6) |
| Extra-Nuclear | * | 1.3/0.9 | 5.6 (-1, -6, 6)/5.3 (3, -6, 2) |
| Third Ventricle | * | 0.1/0.6 | 5.2 (-1, -6, 2)/4.7 (1, -9, -1) |
| Lateral Ventricle | * | 0.1/0.1 | 4.7 (-3, -1, 4)/4.2 (6, -2, 11) |
| Cerebellar Lingual | * | 0.1/0.4 | 3.8 (-1, -46, -15)/4.0 (1, -46, -13) |
| Sub-Gyral | * | 0.2/0.1 | 3.9 (-24, -42, 0)/3.8 (25, -42, 1) |
| Declive | * | 0.3/0.3 | 3.9 (0, -66, -14)/3.6 (0, -63, -17) |
| Declive of Vermis | * | 0.1/0.1 | 3.5 (-3, -69, -14)/3.7 (0, -69, -17) |
| Culmen | * | 0.2/0.3 | 3.7 (-1, -46, -10)/3.7 (4, -46, -10) |
| Parahippocampal Gyrus | * | 0.2/0.1 | 3.6 (-25, -39, -3)/3.5 (25, -39, 3) |
| Culmen of Vermis | * | 0.1/0.1 | 3.5 (0, -66, -9)/3.6 (3, -63, -9) |
| ***tIV-RS8*** | | | |
| Inferior Occipital Gyrus | 17, 18, 19 | 1.3/1.7 | 7.8 (-37, -84, -11)/12.7 (24, -92, -8) |
| Lingual Gyrus | 17, 18 | 1.0/2.2 | 6.2 (-15, -94, -10)/12.6 (19, -94, -7) |
| Fourth Ventricle | * | 0.1/0.2 | 6.4 (-3, -43, -19)/9.3 (0, -40, -22) |

| | | | |
|---|---|---|---|
| Fusiform Gyrus | 18, 19, 37 | 0.4/1.1 | 6.8 (-28, -91, -11)/8.9 (24, -91, -12) |
| Middle Occipital Gyrus | 18, 19, 37 | 0.4/1.3 | 6.0 (-46, -73, -10)/8.3 (42, -79, -10) |
| Cuneus | 17, 18 | 0.1/0.9 | 3.7 (-3, -99, 2)/7.4 (7, -96, 4) |
| Parahippocampal Gyrus | 28, 34 | 0.5/0.3 | 6.9 (-12, -7, -17)/6.5 (12, -5, -16) |
| Third Ventricle | * | 0.0/0.3 | -999.0 (0, 0, 0)/5.6 (0, -23, -1) |
| Uncus | 28, 34 | 0.3/0.1 | 5.4 (-15, -7, -20)/3.6 (15, 1, -19) |
| Lateral Ventricle | * | 0.3/0.4 | 5.1 (-3, -1, 17)/4.6 (3, 2, 12) |
| Superior Parietal Lobule | 7 | 0.1/0.3 | 4.3 (-40, -53, 57)/4.6 (33, -52, 59) |
| Postcentral Gyrus | 1, 2, 3, 40 | 0.0/0.5 | -999.0 (0, 0, 0)/4.6 (43, -32, 62) |
| Nodule | * | 0.0/0.1 | -999.0 (0, 0, 0)/4.6 (0, -46, -27) |
| Culmen | * | 0.3/0.1 | 4.5 (-6, -40, -22)/4.2 (6, -40, -22) |
| Tuber | * | 0.0/0.1 | -999.0 (0, 0, 0)/4.3 (52, -49, -22) |
| Superior Frontal Gyrus | 6, 8, 9 | 0.4/0.1 | 4.3 (-16, 38, 52)/3.5 (15, 33, 56) |
| Inferior Temporal Gyrus | * | 0.0/0.1 | -999.0 (0, 0, 0)/4.1 (52, -57, -11) |
| Inferior Parietal Lobule | 40 | 0.2/0.4 | 3.9 (-48, -42, 56)/4.1 (45, -37, 56) |
| Caudate | * | 0.1/0.0 | 3.9 (-6, 0, 10)/-999.0 (0, 0, 0) |
| Medial Frontal Gyrus | 10 | 0.1/0.0 | 3.8 (-16, 62, -3)/-999.0 (0, 0, 0) |
| Inferior Frontal Gyrus | * | 0.1/0.0 | 3.7 (-37, 9, -14)/-999.0 (0, 0, 0) |
| Middle Frontal Gyrus | 8 | 0.1/0.0 | 3.6 (-28, 39, 44)/-999.0 (0, 0, 0) |
| Extra-Nuclear | * | 0.1/0.1 | 3.5 (-6, -2, 13)/3.5 (18, -32, 25) |

Voxels with z-values greater than 3.5 were matched with the Talairach Daemon database to obtain anatomical labels and converted into MNI space. For each hemisphere, the maximum z-value and MNI coordinate are provided. The volume of voxels in each area is given in cubic centimetres (cc) (*) = area not recognized by standard BA atlas.
